# Confounder-Adjusted Plasma Proteomics Identify Robust Protein Signatures in Sepsis Across Two Independent Perioperative Control Cohorts

**DOI:** 10.64898/2026.09.18.26363373

**Authors:** Maike Weber, Andrea Witowski, Malte Bayer, Frederick Krause, Robin Grugel, Katrin Maria Willemsen, Britta Westhus, Kristin Fuchs, Birgit Zuelch, Samuel Busch, Tim Rahmel, Hartmuth Nowak, Björn Koos, Katharina Rump, Dominik Ziehe, Martin Eisenacher, Michael Adamzik, Barbara Sitek, Thilo Bracht

## Abstract

**Background:** Sepsis is the primary driver of mortality in intensive care. Personalized medicine requires an accurate understanding of the syndrome, and plasma proteomics is increasingly used to characterize its molecular alterations. Comorbidities, demographics and postoperative sterile inflammation complicate this task, and proteins associated with them may be mistaken as sepsis-specific.

**Methods:** We compared 343 sepsis patients with two independent perioperative control cohorts (n = 73 and n = 75) sampled before and after surgery. Plasma was analyzed by mass spectrometry, and analyzed using linear models adjusted for age, sex and cohort-specific comorbidities. Propensity score matching was used to characterize renal comorbidities and Elastic Net classifiers were trained per cohort and time point.

**Results:** We show that renal comorbidities had the strongest impact on the plasma proteome and affected sepsis-associated proteins including Cystatin-C and Beta-2-microglobulin. Eleven proteins were robustly associated with sepsis, among them established acute phase proteins and the less well characterized Beta-1,4-galactosyltransferase 1. Feature importance analysis confirmed this core set and identified further proteins that contributed in a cohort- and time point-specific manner. Classifiers performed very well except against control patients at day 3 after surgery, when sterile inflammation peaked.

**Conclusions:** Sepsis, comorbidities and sterile inflammation may alter the same acute phase and renal proteins, differing in magnitude rather than in the type of response. We disentangled these influences and report a robust set of sepsis-associated proteins together with confounder-associated proteins, providing a basis for future studies and the development of diagnostic and therapeutic strategies.

**Plain Language Summary:** Sepsis is a life-threatening response to infection and a leading cause of hospital death. Precision medicine requires an understanding of its molecular basis. Plasma proteins provide access to it, but are also influenced by comorbidities, age and sterile inflammation following surgery. We analyzed the plasma proteome of 343 sepsis patients and 148 surgical patients without infection, sampled before and after surgery, adjusting for age, sex and comorbidities. Eleven proteins were robustly associated with sepsis. Impaired kidney function had the strongest influence, and samples taken at the peak of sterile inflammation were hardest to distinguish from sepsis. Our results disentangle the influence of sepsis, comorbidities and sterile inflammation on the plasma proteome, paving the way for novel diagnostic and therapeutic strategies.

## Introduction

Sepsis remains one of the leading causes of death worldwide and is the primary driver of mortality in intensive care units, despite decades of increasing research efforts. To date, effective treatment is still limited to antimicrobial therapy targeting the causative pathogen and hemodynamic stabilization through fluid resuscitation and vasopressor support [1]. Other therapeutic approaches failed due to the heterogeneity of the syndrome, and personalized medicine is widely regarded as a key strategy to reduce sepsis mortality.

Developing targeted therapeutic strategies requires a precise understanding of the molecular changes underlying the syndrome [2,3]. Related etiologies must also be studied to distinguish sepsis-specific alterations from those occurring in other conditions, such as sterile inflammation. Furthermore, sepsis must be reliably diagnosed at the earliest possible time point, as treatment delay is associated with poor outcomes [4].

To address these problems, high-throughput proteomic approaches are increasingly employed to characterize the molecular landscape of sepsis and to compare patients with hospital controls or healthy donors. The preferred sample is serum or plasma, owing to their comparatively straightforward availability and because the bloodstream represents the primary compartment affected in sepsis. Recently, a large-scale study characterized the plasma proteome of sepsis patients in comparison to healthy volunteers, postoperative patients, and non-infected ICU controls, identifying acute-phase, complement, and lipoprotein-related proteins as core features of the sepsis response [5]. Other studies have proposed a plasma protein panel to distinguish sepsis from sterile inflammation [6] and compared serum from sepsis patients and healthy controls to predict outcome within the sepsis cohort [7]. However, the influence of comorbidities and demographic confounders such as age and sex on the composition of the plasma proteome was frequently overlooked, and proteins associated with confounders might have been mistaken as sepsis-associated. Another important issue when comparing sepsis and control patients is confounding by sterile inflammation induced by major surgery when ICU or severely affected ward patients serve as controls. Acute-phase proteins and even canonical markers such as C-reactive protein (CRP) are upregulated in the absence of infection and have the potential to interfere with diagnostic biomarkers.

In the present study, we addressed these hurdles by analyzing sepsis patients alongside two perioperative control cohorts, sampled before and after surgery (Figure 1). Furthermore, we systematically evaluated and adjusted for the influence of comorbidities and demographic factors on the plasma proteome. using linear models, propensity score matching and machine learning. The aim of the study was to identify proteins that robustly discriminate between patients with sepsis and hospital controls, providing a stronger foundation for the development of diagnostic and therapeutic strategies.

**Figure 1.**
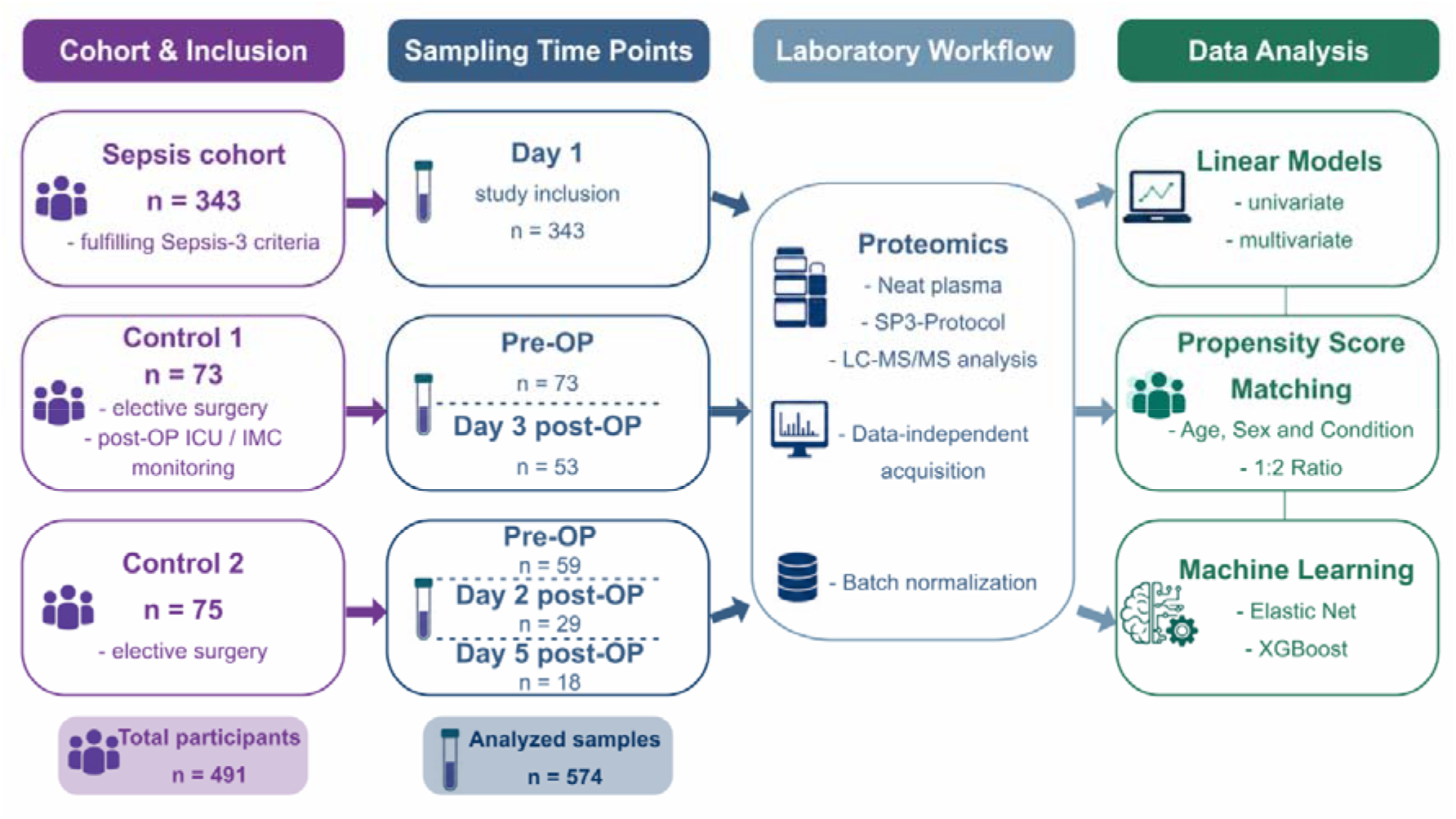
Schematic representation of the study cohort and the analytical workflow.

## Results

### Clinical characteristics differ between sepsis patients and control cohorts

The study cohort comprised 343 patients with sepsis, 73 hospitalized controls undergoing planned surgery who required postoperative intensive or intermediate care monitoring (Control 1), and 75 hospitalized controls undergoing planned surgery who did not require such monitoring (Control 2; Table 1). Sepsis patients and Control 1 were of comparable age, whereas Control 2 patients were significantly older (p < 0.001). Sex distribution was similar across groups. Among septic patients, the median SOFA score was 8 (5–11), with pulmonary (49.9%) and abdominal (22.4%) infections most common. No active infection was present in either control cohort by definition.

**Table 1.** Baseline characteristics of the study cohorts.

| Variable | Level | Sepsis<br>(n = 343) | Control 1<br>(n = 73) | Control 2<br>(n = 75) | p-value | Posthoc-p-values |  |  |
| --- | --- | --- | --- | --- | --- | --- | --- | --- |
|  |  |  |  |  |  | Sepsis vs<br>Control 1 | Sepsis vs Con-<br>trol 2 | Control 1 vs Con-<br>trol 2 |
| Age, years |  | 66.0<br>[56.0–75.0] | 65.0<br>[57.0–76.0] | 77.0<br>[70.0–83.0] | **1.22E-09** | 1 | **1.03E-09** | **1.88E-06** |
| Sex, female |  | 138 (40.2%) | 35 (47.9%) | 33 (45.2%) | 0.395 | 0.7224 | 1 | 1 |
| BMI, kg/m<sup>2</sup> |  | 26.9<br>[24.2–30.6] | 24.7<br>[23.2–29.4] | 28.7<br>[25.2–32.2] | **0.001** | **0.0283** | 0.1138 | **0.0006** |
| SOFA score |  | 8.0 [5.0–11.0] | 3.0 [1.0–6.0] |  |  |  |  |  |
| Infection focus |  | N = 343 | N = 73 |  |  |  |  |  |
|  | None | 8 (2.3%) | 73 (100.0%) | 75 (100.0%) |  |  |  |  |
|  | Abdomen | 77 (22.4%) |  |  |  |  |  |  |
|  | Bloodstream | 40 (11.7%) |  |  |  |  |  |  |
|  | Urinary tract | 26 (7.6%) |  |  |  |  |  |  |
|  | Lung | 171 (49.9%) |  |  |  |  |  |  |
|  | CNS | 10 (2.9%) |  |  |  |  |  |  |
|  | Other | 11 (3.2%) |  |  |  |  |  |  |
| Surgery Duration, minutes |  | NA | 198.0<br>[140.5–354.5] | 103.5<br>[81.0–122.5] | 1.08E-11 |  |  |  |
| Malignancy |  | 101 (29.5%) | 58 (79.5%) | 19 (25.3%) | **0.0005** | **9.45E-15** | 1 | **7.23E-11** |
| Transplant |  | 47 (13.7%) | 1 (1.4%) | 1 (1.3%) | **0.0005** | **0.0030** | **0.0030** | 1 |
| Diabetes mellitus |  | 104 (30.3%) | 9 (12.3%) | 14 (18.7%) | **0.0015** | **0.0040** | 0.1421 | 1 |
| Nicotine use |  | 64 (18.7%) | 26 (35.6%) | 5 (6.7%) | **0.0005** | **0.0079** | **0.0289** | **4.06E-05** |
| Chronic kidney disease |  | 64 (18.7%) | 4 (5.5%) | 21 (28.0%) | **0.0010** | **0.0142** | 0.2436 | **0.0009** |
| Other lung disease |  | 44 (12.8%) | 11 (15.1%) | 26 (34.7%) | **0.0005** | 1 | **6.96E-05** | **0.0229** |
| Obesity |  | 74 (21.8%) | 15 (20.5%) | 29 (38.7%) | **0.0095** | 1 | **0.0093** | 0.0586 |
| Cardiovascular disease |  | 90 (26.3%) | 23 (31.5%) | 28 (37.3%) | 0.1394 | 1 | 0.1967 | 1 |
| COPD |  | 51 (14.9%) | 12 (16.4%) | 8 (10.7%) | 0.5607 | 1 | 1 | 1 |
| Alcohol abuse |  | 28 (8.2%) | 8 (11.0%) | 7 (9.3%) | 0.6422 | 1 | 1 | 1 |
| Hypertension |  | 230 (67.1%) | 46 (63.0%) | 51 (68.0%) | 0.7761 | 1 | 1 | 1 |
| Dialysis |  | 34 (10.0%) | 0 (0.0%) | 0 (0.0%) | **0.0005** |  |  |  |

Comorbidity profiles differed between the groups. Malignancy was substantially more common in Control 1, while kidney transplantation history and dialysis dependency were largely confined to the sepsis cohort. Diabetes mellitus was most frequent among septic patients, whereas chronic kidney disease (CKD) and cardiovascular disease were most prevalent in Control 2 (all p < 0.05, Table 1). Lifestyle-associated risk factors also varied: nicotine abuse was most common in Control 1, while alcohol abuse was more frequent in Control 2 (both p < 0.05). Body mass index (BMI) differed significantly between groups (p = 0.001), whereas obesity, hypertension, COPD, and other chronic lung diseases did not.

Surgical procedure types also differed between control cohorts, with joint replacement and spinal surgery more common in Control 2 and visceral and oral/maxillofacial surgery more common in Control 1 (all p < 0.05, Supplementary Table 1). Cranial neurosurgery was performed only in Control 1 (38%), per the study protocol. Surgery duration differed significantly between Control 1 and 2 (198.0 (140.5–354.5) minutes vs. 103.5 (81.0–122.5) minutes, p < 0.001). Overall, the sepsis cohort was characterized by organ dysfunction and a higher burden of kidney transplantation, diabetes, and dialysis dependency, while Control 1 showed enrichment for malignancies and Control 2 was older with greater cardiovascular and renal comorbidity.

### Linear models reveal the impact of renal conditions on the plasma proteome

Univariate linear models were used to analyze the influence of demographic factors and comorbidities on the plasma proteome. The models were fitted individually for Control 1 and Control 2 (pre-OP time points), sepsis patients, and the aforementioned cohorts combined (Figure 2a). While for sepsis patients an influence of the renal conditions (CKD, transplant and dialysis) was observed, the combined analysis primarily revealed a significant bias driven by the strongly imbalanced comorbidities malignancy, transplant and dialysis (Table 1). As these comorbidities were specific to either the sepsis or the control cohorts, the significantly associated proteins were primarily related to the differences between these conditions. Thus, the univariate analysis flagged the imbalanced comorbidities as problematic for the multivariate analysis. Here, we first evaluated the collinearity of all covariates. We found a moderate correlation among the renal conditions transplant, dialysis and CKD. As expected, BMI correlated with obesity, and a weak association between alcohol and nicotine was observed (Figure 2b). Within the sepsis cohort, we also observed a weak correlation of COPD with nicotine abuse, as well as of hypertension with BMI (Supplementary Figure 1). The correlation among comorbidities did not require exclusion of individual parameters; however, the imbalance of transplant and dialysis prevented their inclusion in the multivariate models. Accordingly, only CKD was included in the model comparing Control 1 with sepsis patients. Individual models were used for both control cohorts, adjusted for the demographic confounders age and sex, as well as for the comorbidities that showed differential prevalence between the compared cohorts (Table 1). Both multivariate models showed the most significant associations with the primary variable (i.e. condition, sepsis vs. control, Supplementary Tables 6-10). Interestingly, no significant associations were found for most of the imbalanced comorbidities in the multivariate analysis; however, significant associations were found for age, sex and chronic kidney disease.

**Figure 2.**
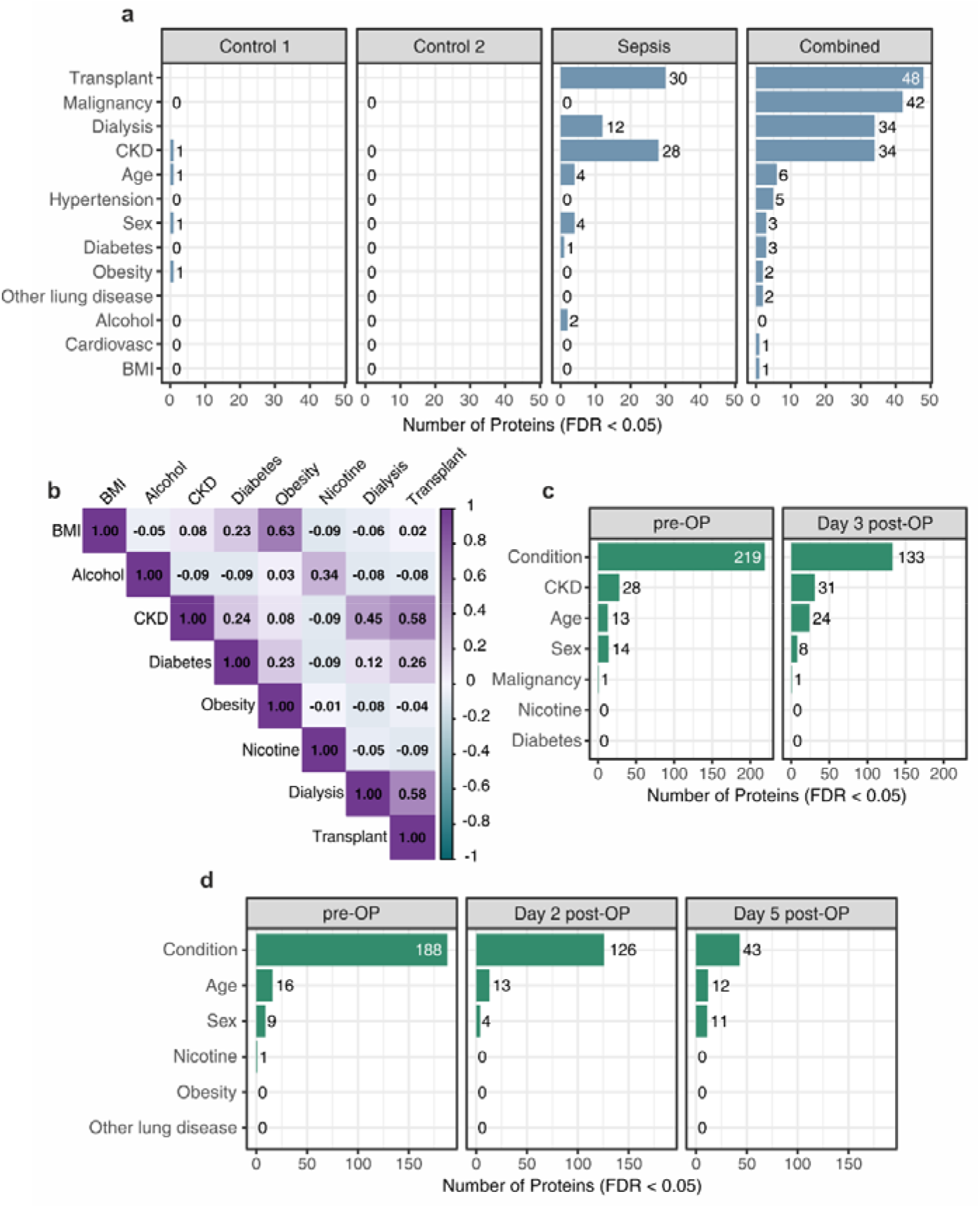
Analysis of the influence of comorbidities by linear models. **a)** All comorbidities were tested using univariate linear models, applied separately to Control 1 (pre-OP), Control 2 (pre-OP), the sepsis cohort, and the combination of all three. Bars represent the numbers of proteins significantly associated with the respective comorbidities (only comorbidities with significant associations are shown). **b)** Pearson correlation matrix for all comorbidities as well as age, sex and BMI (calculated on the combined cohort; only terms with at least one correlation > |0.25| displayed). **c)** Multivariate analysis considering condition (sepsis vs. control) adjusted for the displayed covariates. The model was applied for differentiation of sepsis patients and Control 1 at two time points. **d)** Multivariate analysis used to distinguish between sepsis patients and Control 2 at three time points. All p-values adjusted according to the Benjamini-Hochberg procedure.

### An independent matching approach confirms renal-associated proteome alterations

To further investigate the influence of comorbidities that were strongly imbalanced between the sepsis and the control cohorts, we applied propensity score matching as an approach independent of the linear models. For each comorbidity, patients carrying it were compared to patients not carrying it. For every carrier, two non-carriers that were matched for age, sex and condition (sepsis or control) were selected from the combined patient pool. The resulting sub-cohorts were therefore balanced for these variables and were compared using t-tests. Minor differences were observed for diabetes and obesity (2 and 5 significant proteins, respectively; Supplementary Table 11), while the influence of renal conditions on the plasma proteome was confirmed (Figure 3a). The known marker of kidney function Cystatin-C (CST3) for instance, was found to be higher abundant in all three comparisons as well as Alpha-1-microglobulin (AMBP), which is known for accumulation in plasma in chronic kidney disease patients. The results from linear models and propensity score matching were largely in agreement, thereby substantiating the observations made by both approaches (Figure 3b). Many of the observed proteins are small and freely filtered by the healthy kidney and accumulate in the plasma when kidney function is impaired (Table 2). The COL18A1 gene also codes for Endostatin, a potent inhibitor of angiogenesis of about 20 kDa mass.

**Figure 3.**
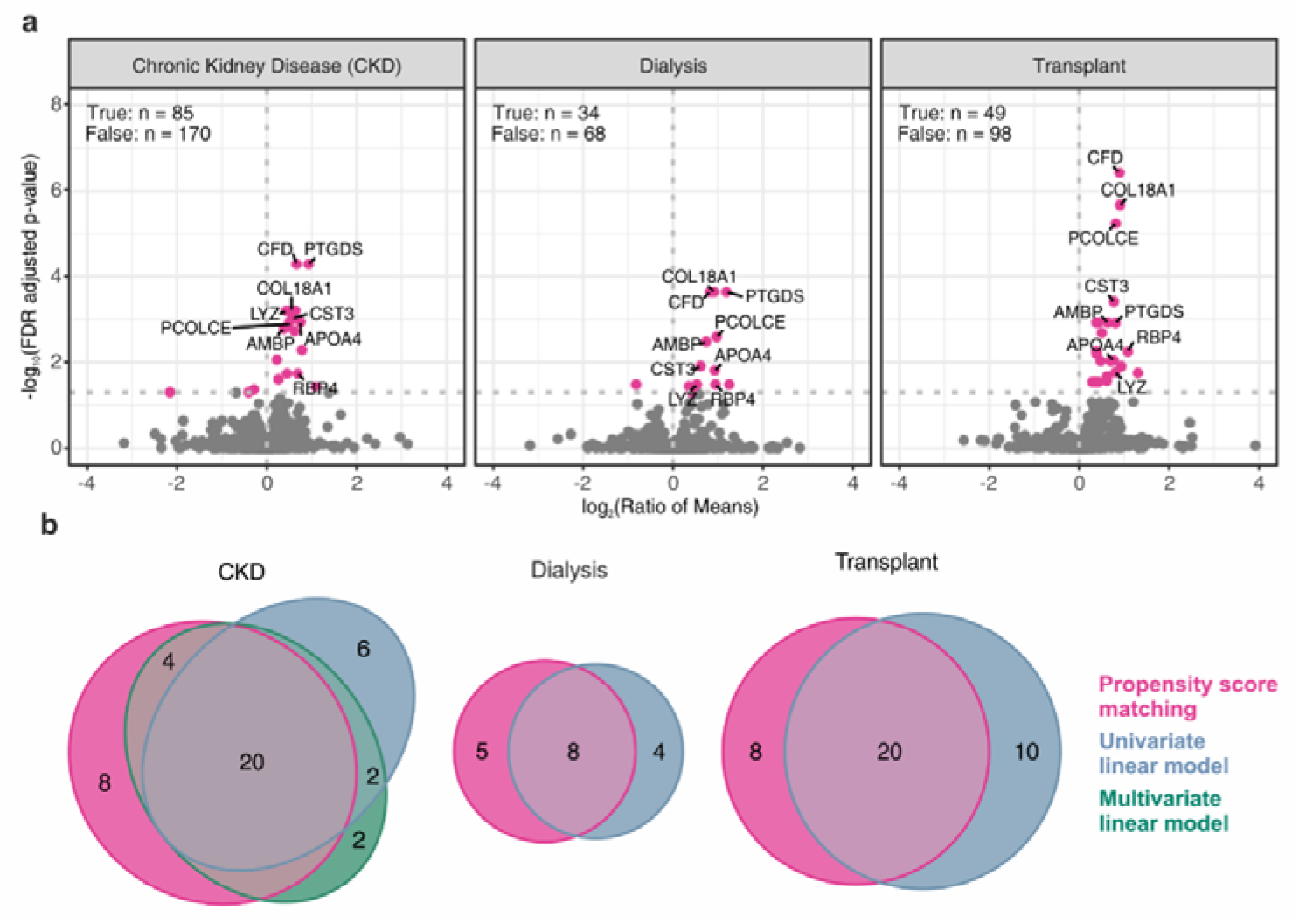
Propensity score matching and comparison with linear models. **a)** For each comorbidity, patients carrying it were matched to two patients not carrying it, based on age, sex and condition (sepsis or control). Patient numbers are displayed in the upper left corner of the plots. Volcano plots represent the comparison of carriers and non-carriers by t-test (p-values adjusted using the Benjamini–Hochberg method; ratios of means calculated as carriers/non-carriers). Proteins found significant in all three comparisons are labeled with gene names. **b)** Euler diagrams representing the overlap of significant proteins identified by linear models (univariate models fitted only on sepsis patients, multivariate model for CKD fitted for the comparison with Control 1) and by analysis of propensity score matched sub-cohorts.

**Table 2.**
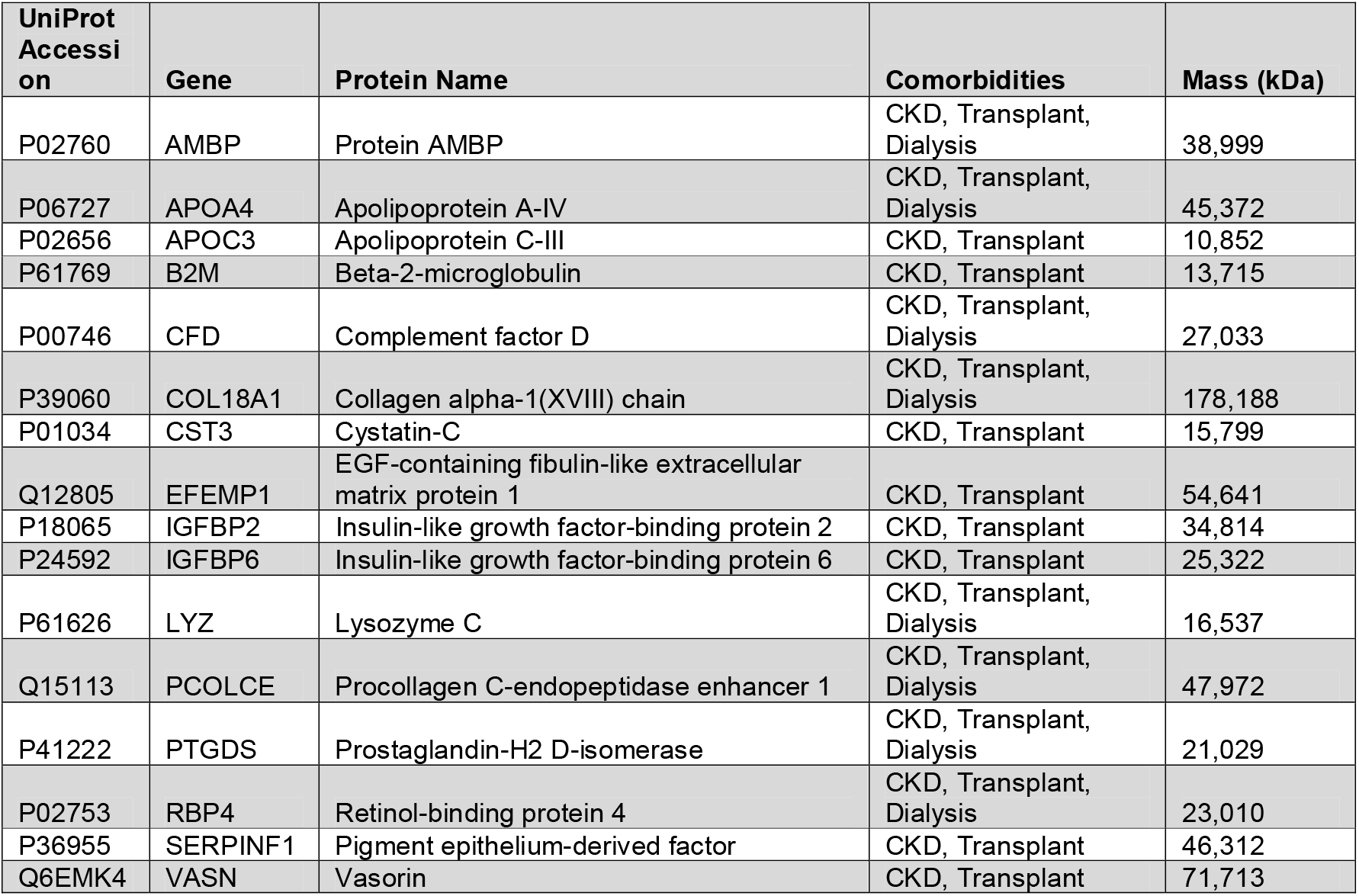
Proteins consistently associated with renal comorbidities. Displayed proteins were found in the overlap of linear models and propensity score matching for at least two of three renal comorbidities.

| UniProt Accession | Gene | Protein Name | Comorbidities | Mass (kDa) |
| --- | --- | --- | --- | --- |
| P02760 | AMBP | Protein AMBP | CKD, Transplant, Dialysis | 38,999 |
| P06727 | APOA4 | Apolipoprotein A-IV | CKD, Transplant, Dialysis | 45,372 |
| P02656 | APOC3 | Apolipoprotein C-III | CKD, Transplant | 10,852 |
| P61769 | B2M | Beta-2-microglobulin | CKD, Transplant | 13,715 |
| P00746 | CFD | Complement factor D | CKD, Transplant, Dialysis | 27,033 |
| P39060 | COL18A1 | Collagen alpha-1(XVIII) chain | CKD, Transplant, Dialysis | 178,188 |
| P01034 | CST3 | Cystatin-C | CKD, Transplant | 15,799 |
| Q12805 | EFEMP1 | EGF-containing fibulin-like extracellular matrix protein 1 | CKD, Transplant | 54,641 |
| P18065 | IGFBP2 | Insulin-like growth factor-binding protein 2 | CKD, Transplant | 34,814 |
| P24592 | IGFBP6 | Insulin-like growth factor-binding protein 6 | CKD, Transplant | 25,322 |
| P61626 | LYZ | Lysozyme C | CKD, Transplant, Dialysis | 16,537 |
| Q15113 | PCOLCE | Procollagen C-endopeptidase enhancer 1 | CKD, Transplant, Dialysis | 47,972 |
| P41222 | PTGDS | Prostaglandin-H2 D-isomerase | CKD, Transplant, Dialysis | 21,029 |
| P02753 | RBP4 | Retinol-binding protein 4 | CKD, Transplant, Dialysis | 23,010 |
| P36955 | SERPINF1 | Pigment epithelium-derived factor | CKD, Transplant | 46,312 |
| Q6EMK4 | VASN | Vasorin | CKD, Transplant | 71,713 |

Nine of 13 quantified COL18A1 peptides overlapped with the sequence of Endostatin suggesting that it is likely that endostatin was the primary protein measured (Supplementary Figure 2).

### Multivariate linear models reveal plasma proteome alterations in sepsis

The previously developed multivariate models were used to analyze plasma proteome differences between patients with sepsis and control patients, adjusting the cohort-specific confounders. Both models were adjusted for age and sex; the model for Control 1 was additionally adjusted for CKD, malignancy, nicotine abuse and diabetes, the model for Control 2 for nicotine abuse, obesity and other lung diseases. Proteins with an adjusted p-value < 0.05 and a ratio of means ≥ 1.5 or ≤ 0.67 were considered differentially abundant.

Most differential proteins were found for the comparison with Control 1 pre-OP (Figure 4a), with fewer significant proteins detected at day 3 post-OP. This likely reflected proteome changes that can be attributed to the sterile inflammation occurring after surgery as several acute inflammatory proteins were upregulated post-OP (Figure 4b). For Control 2, the difference between pre- and post-operative time points was less pronounced.

**Figure 4.**
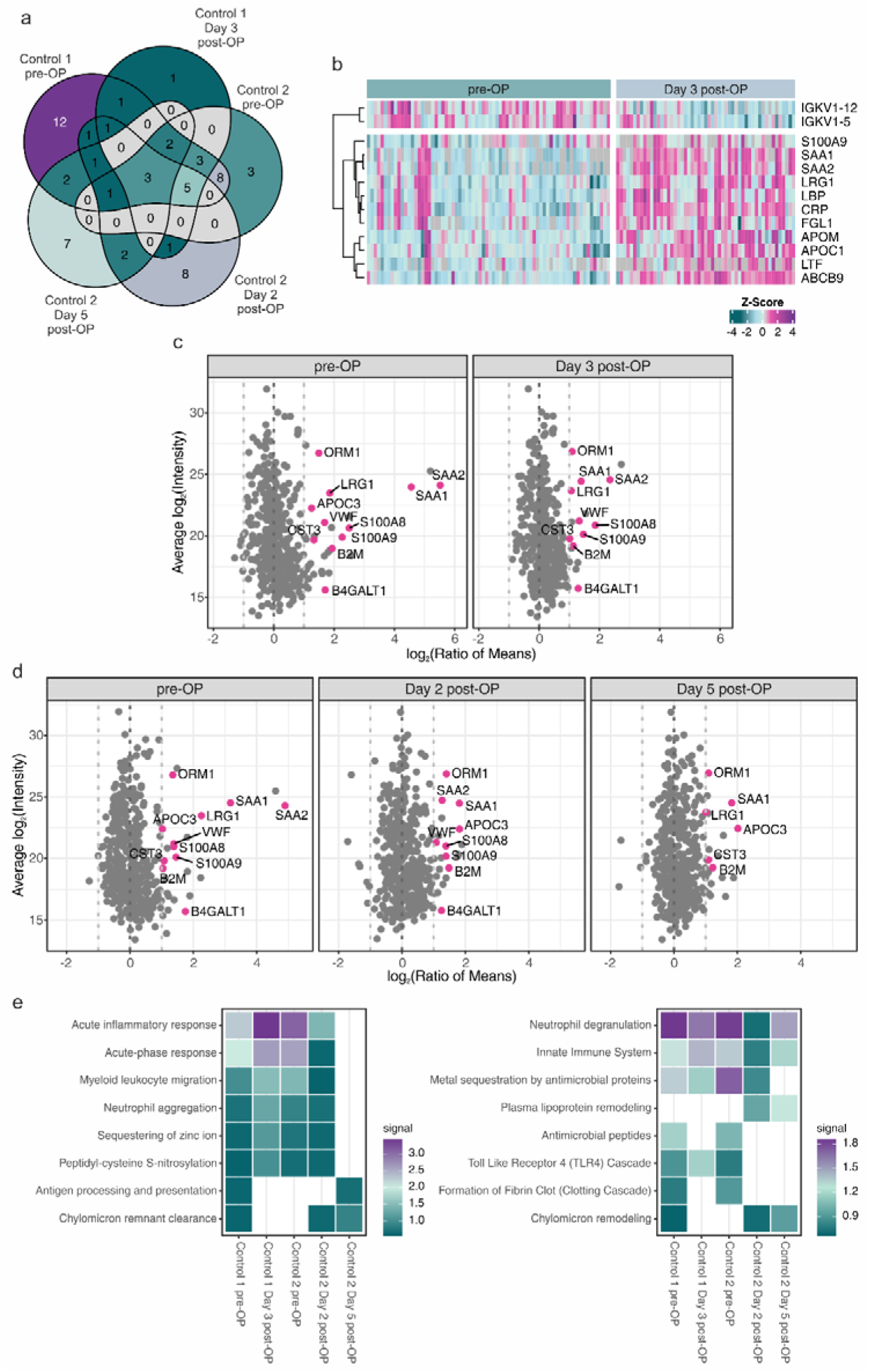
Differential analysis of sepsis and control patients using multivariate linear models. **a)** Venn diagram illustrating the five pairwise comparisons and the respective numbers of significantly differentially abundant proteins (adjusted p-value < 0.05, absolute ratio of means > 1.5) as well as the overlaps of the corresponding proteins. **b)** Plasma proteome changes between pre- and post-OP time points for Control 1. Displayed proteins were found to be significantly regulated between the two time points (t-test, adjusted p-value < 0.05, absolute ratio of means > 1.5). **c)** Comparison of sepsis patients with Control 1 using a multivariate linear model correcting for age, sex and four comorbidities illustrated as MA plots (to account for systematic differences in p-values due to different statistical power). Proteins that were found differential in at least four of five comparisons in **c)** and **d)** are highlighted and labeled with gene names. **d)** Comparison of sepsis patients with Control 2 using a multivariate linear model correcting for age, sex and three comorbidities. **e)** Functional enrichment analysis was carried out using STRING. Significantly differential abundant proteins were analyzed separately for each comparison and eight selected significantly enriched GO biological processes (left panel) and REACTOME pathways (right panel) are displayed.

The numbers of significant proteins varied considerably across the comparisons (Figure 4a, Supplementary Table 12), but for all comparisons, the majority of significant proteins were upregulated in sepsis patients (Figure 4c and d). The corresponding proteins were mostly related to acute inflammatory and immune processes, but lipoprotein metabolism, which is known to be related to sepsis severity [8], was also affected (Figure 4e). Eleven proteins overlapped in at least four of the five comparisons and were therefore considered robustly specific to the sepsis plasma proteome (Table 3). Several of these proteins are well documented in sepsis but were also found to be regulated in sterile inflammation (Figure 4b), rendering the difference quantitative rather than qualitative.

**Table 3.**
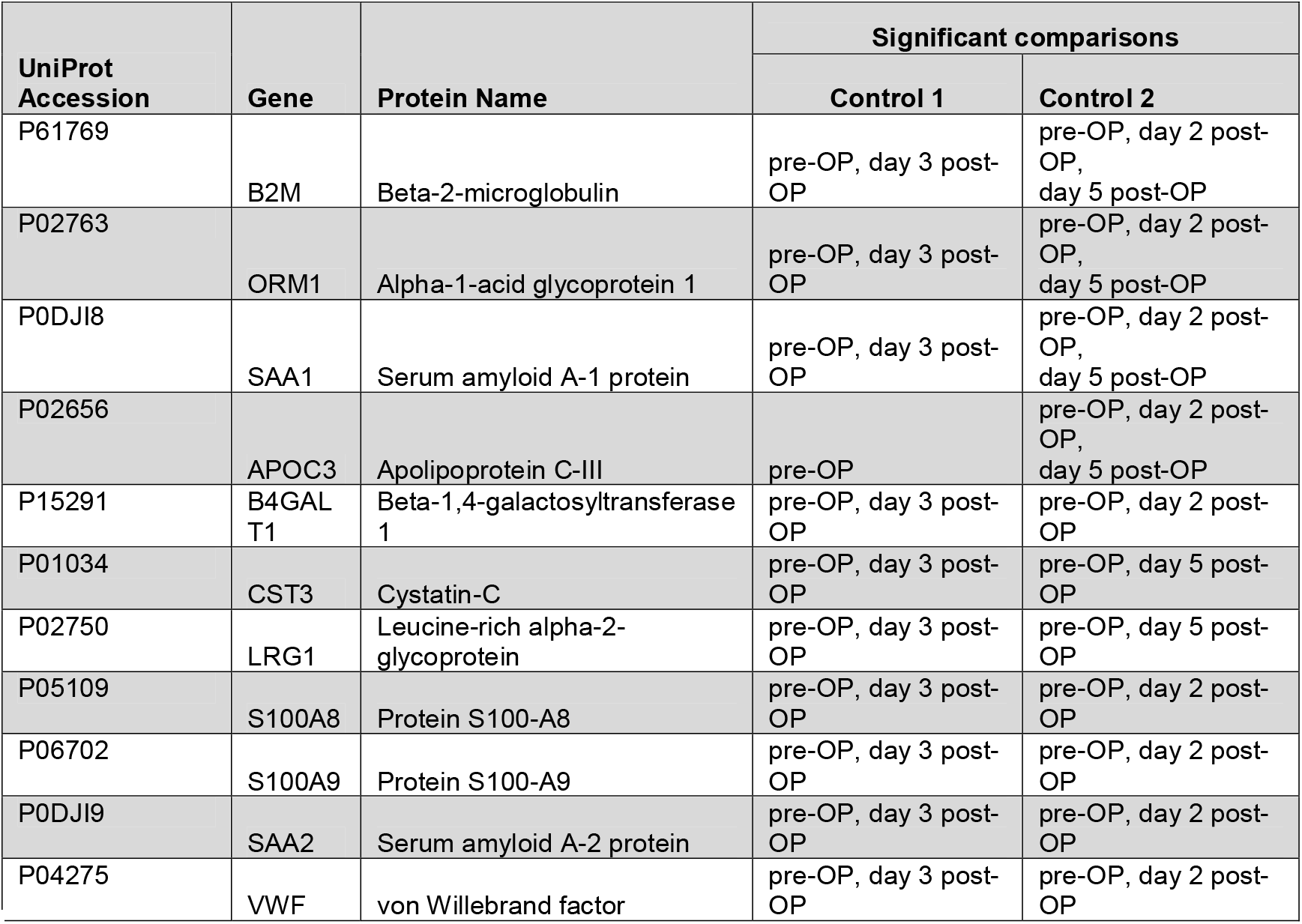
Robustly sepsis-associated proteins. Displayed proteins were found significant between sepsis and control patients in at least four of five analyzed comparisons and are highlighted in Figure 4 c) and d).

Interestingly, the sepsis-associated proteins CST3 and Beta-2-microglobulin (B2M) were also found to be related to renal conditions, suggesting increased renal damage in sepsis patients.

To assess whether these alterations reflected the unequal distribution of renal comorbidities between the cohorts, the five comparisons were repeated after excluding all sepsis patients with kidney transplantation or dialysis dependency (Supplementary Figure 6). Effect sizes were nearly identical in both analyses and the numbers of differentially abundant proteins changed only marginally. All eleven robustly sepsis-associated proteins retained significance in at least four of the five comparisons; only CST3 lost significance in the comparison with Control 2 at day 2 post-OP.

Regarding the investigated confounders, we found several proteins to be significantly associated with age and sex. Following the same filter logic as for sepsis vs. control, we considered proteins significant in at least four of five comparisons to be robustly associated with the respective covariates. For sex, we found five robust proteins, of which Pregnancy zone protein (PZP), Sex hormone-binding globulin (SHBG), as well as Adiponectin (ADIPOQ) can be considered positive controls, as their well-documented sex-related abundance confirmed the validity of our analytical approach [9,10] (Supplementary Table 13). For age, the results were more difficult to interpret as several of the associated proteins were also found to be linked to renal function (CST3, CFD, COL18A1) and sex (SHBG; Supplementary Table 14), indicating collinearity between these confounders. Notably, several age-associated proteins belonged to the extracellular matrix, a subset of which are typically located in the synovium (e.g. Cartilage acidic protein 1, CRTAC1).

### Machine learning models differentiate sepsis from controls

Four binary classifiers were trained to distinguish sepsis patients from controls. Sepsis against Control 1 at the pre-OP and day 3 post-OP time points, and against Control 2 at pre-OP and day 2 post-OP (no model was trained for Control 2 at day 5 post-OP due to insufficient patient numbers). The control group was defined as the positive class and all reported metrics represent means across repeated cross-validation (Figure 5a). Overall, the model performance was very good and comparable across three of the four models for both model types (Elastic Net and XGBoost). Only the differentiation between sepsis patients and Control 1 at day 3 post-OP performed substantially worse (Table 4). Sensitivity dropped from 0.88 to 0.58, indicating that postoperative controls were frequently classified as sepsis, while specificity was less affected (0.96 to 0.90; Supplementary Figure 3). For Control 2, no such decline was observed and performance was comparable at both time points (AUROC 0.98 and 0.99). Taken together, the models illustrate that the plasma proteome carries sufficient information to separate sepsis from perioperative controls. The consistent performance across two independent control cohorts allows an optimistic perspective with regard to the application in independent patient collectives.

**Figure 5.**
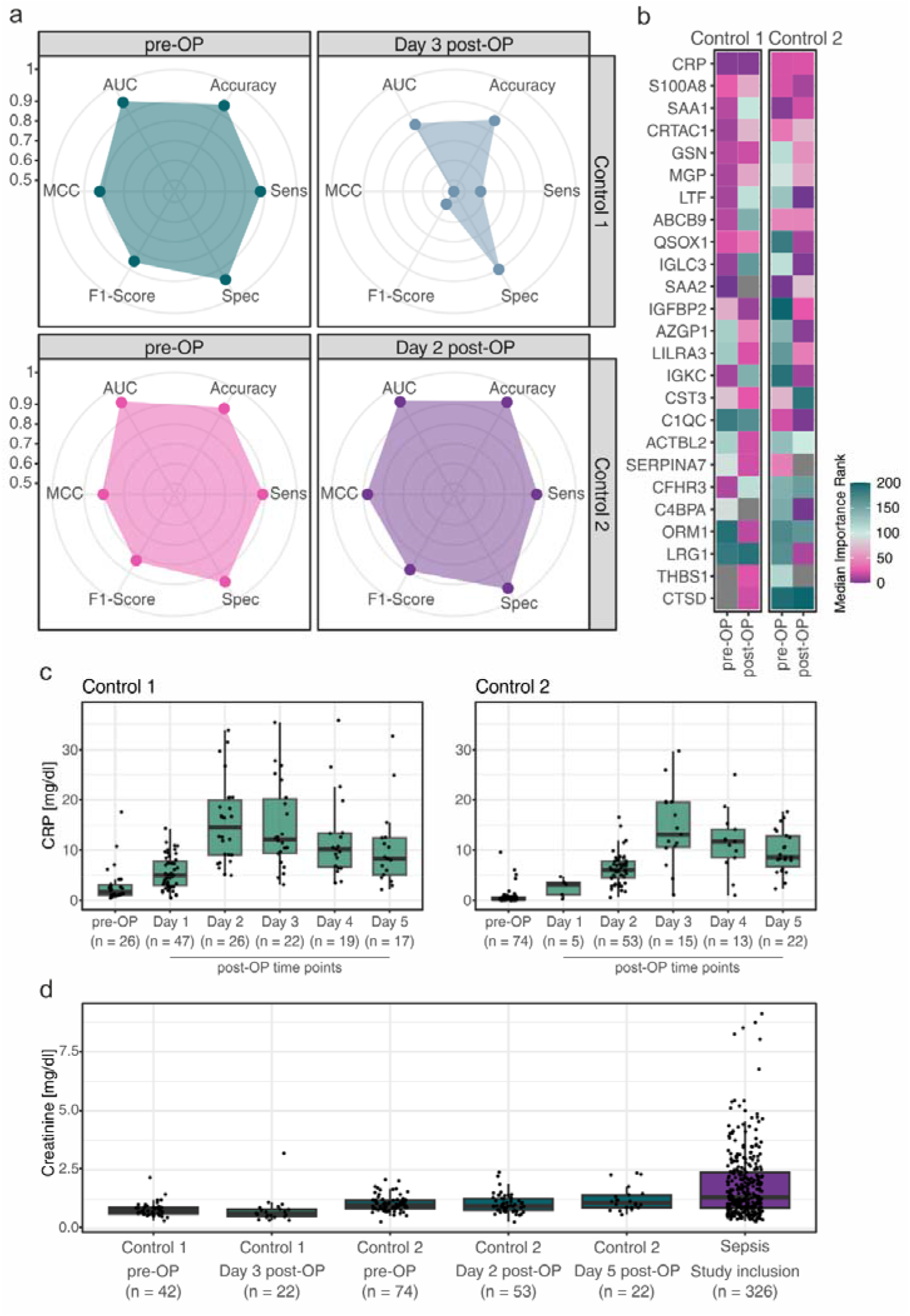
Elastic Net classifiers and feature importance analysis. **a)** Four Elastic Net models were trained to discriminate sepsis patients from Control 1 and 2 at the pre-OP time point, and at day 3 (Control 1) or day 2 (Control 2) post-OP, respectively. Radar plots illustrate the performance metrics of the trained models. **b)** Heatmap showing the 25 most influential features across the four models. For each model, the ten features with the lowest median importance ranks were selected and the cumulative list was trimmed to the top 25 proteins for readability. **c)** C-reactive protein (CRP) concentrations from clinical routine measurements for both control cohorts at the pre-operative and all available postoperative time points; the number of patients is displayed below each time point. **d)** Creatinine concentrations from clinical routine measurements at the time points analyzed in this study.

**Table 4.** Mean performance metrics (standard deviation) of the trained elastic net models.

|  | Control cohort 1 |  | Control cohort 2 |  |
| --- | --- | --- | --- | --- |
|  | pre-OP | Day 3 post-OP | pre-OP | Day 2 post-OP |
| **Accuracy** | 0.95 (± 0.02) | 0.86 (± 0.03) | 0.95 (± 0.02) | 0.98 (± 0.01) |
| **Sensitivity** | 0.88 (± 0.06) | 0.58 (± 0.12) | 0.89 (± 0.08) | 0.86 (± 0.12) |
| **Specificity** | 0.96 (± 0.02) | 0.90 (± 0.03) | 0.96 (± 0.02) | 0.99 (± 0.01) |
| **F1-Score** | 0.85 (± 0.04) | 0.52 (± 0.08) | 0.83 (± 0.06) | 0.89 (± 0.08) |
| **MCC** | 0.82 (± 0.05) | 0.44 (± 0.09) | 0.80 (± 0.07) | 0.88 (± 0.09) |
| **AUROC** | 0.96 (± 0.02) | 0.83 (± 0.05) | 0.98 (± 0.01) | 0.99 (± 0.03) |

Since the Elastic Net models outperformed the XGBoost models in three of four cases based on mean sensitivity (Supplementary Table 15), only the Elastic Net models were used for feature importance analysis. Features were ranked by their median SHAP value across the cross-validation repetitions, with low ranks indicating high importance (Figure 5b, Supplementary Table 16). The established clinical marker CRP was the most consistently important feature and several further prominent features overlapped with the proteins robustly associated with sepsis in the linear models, thereby substantiating their discriminative value. These included the acute phase proteins Serum amyloid A-1 and A-2 (SAA1, SAA2), alpha-1-acid glycoprotein 1 (ORM1), the alarmin S100-A8 and Leucine-rich alpha-2-glycoprotein (LRG1), which is also released by activated neutrophils [11]. The remaining robustly sepsis-associated proteins contributed little to the classifiers, which was expected for regularized models as among correlated proteins, a single representative is retained while redundant features are down-weighted. Consistently important across all four models was Cartilage acidic protein 1 (CRTAC1) a protein that has not been studied in the context of sepsis. Gelsolin (GSN), previously discussed as a biomarker for sepsis [12] and Matrix Gla protein (MGP) were mainly relevant for the discrimination from Control 1.

Beyond a core set of proteins with broad relevance, several features contributed in a cohort- and time point-specific manner. The importance of the SAA proteins decreased at the postoperative time points in both control cohorts, but the decline was considerably more pronounced for Control 1 than for Control 2. Clinical routine data confirmed that this reflected the extent of the postoperative acute phase response at the respective sampling time points (Figure 5c). In Control 1, CRP reached highest concentrations at day 2 and remained elevated at day 3, whereas in Control 2 CRP peaked at day 3. Conversely, ORM1, an acute phase protein with slower kinetics [13], was especially relevant in the Control 1 post-OP comparison. Complement factor H-related protein 3 (CFHR3) showed a low rank only for Control 1 pre-OP, and C4b-binding protein alpha chain (C4BPA) was of high importance exclusively in Control 2 at postoperative day 2, in agreement with the linear model, where it showed the highest significance for the respective comparison (Supplementary Table 9).

Taken together, a core set of inflammatory proteins consistently drove model performance across cohorts and time points, while additional features contributed in a more context-specific manner, reflecting the distinct clinical trajectories of the individual control groups.

## Discussion

Distinguishing sepsis-specific alterations of the plasma proteome from those driven by comorbidities, demographics, and sterile inflammation remains a major analytical challenge. Here we compared sepsis patients with two independent perioperative control cohorts, adjusting for age, sex, and the comorbidities that differed between the compared groups.

Eleven proteins were robustly associated with sepsis, comprising established markers alongside less extensively characterized proteins. Combining linear models with the analysis of propensity score-matched sub-cohorts revealed markers of renal function with the potential to confound sepsis-associated proteome changes. Machine learning classifiers separated sepsis from controls with very good performance, except for the comparison with postoperative Control 1 patients. Analyzing two control populations at several perioperative time points thus allowed us to isolate a core signature that is robust to the control cohort and the sampling time point.

In the present study, all proteins found to be robustly associated with sepsis by linear models were upregulated compared to controls and had previously been reported in the context of sepsis. Alpha-1-acid glycoprotein 1 (ORM1), Serum amyloid A-2 protein (SAA2) and von Willebrand factor (VWF) were recently reported to be upregulated in sepsis in comparison with non-infectious systemic inflammatory response syndrome [6]. Serum amyloid A-1 protein (SAA1) and Protein S100-A9 (S100A9) were described in comparison to healthy controls [14]. Likewise, Apolipoprotein C-III (APOC3) and Beta-2-microglobulin (B2M) were described to be elevated in comparison to non-septic hospital controls [15] and Leucine-rich alpha-2-glycoprotein (LRG1) was reported as upregulated in pediatric sepsis [16]. One protein that was less extensively reported within a sepsis context was the Beta-1,4-galactosyltransferase 1 (B4GALT1), which was found to be significantly upregulated in sepsis in four of five comparisons (Figure 4 c and d). B4GALT1 is a key enzyme in glycan biosynthesis and catalyzes the transfer of galactose onto N-glycans. Recently, B4GALT1 was identified as a regulator of cytotoxic T cell function, enabling tumor immune escape by regulating the glycosylation of the T cell receptor and the immune checkpoint [17,18], which established its relevance in immune regulation. Apart from a report by Mi et al., in which B4GALT1 was differential between sepsis and several control groups [5], the protein has received little attention in this context, and its role in inflammation and sepsis has yet to be clarified.

We found Cystatin-C (CST3), B2M and APOC3 to be robustly upregulated in sepsis but also linked to renal conditions (Table 2). In addition, CST3 was amongst the most important machine learning features (Figure 5b). CST3 and B2M are established markers of renal function, and APOC3 has also been linked to chronic kidney disease [19]. We previously described CST3, B2M and Complement factor D (CFD) to be related to kidney injury in sepsis and showed that their plasma levels correspond to kidney damage, as represented by creatinine [20]. Since kidney transplantation and dialysis dependency were almost exclusively found in the sepsis cohort (Table 1), these findings raise the question whether the observed protein alterations merely reflect the unequal distribution of renal diseases.

Several observations argue against this interpretation. Excluding the patients with kidney transplantation or dialysis dependency left the effect sizes virtually unchanged, and all eleven robustly sepsis-associated proteins remained significant in at least four of the five comparisons (Supplementary Figure 5). Moreover, chronic kidney disease was more prevalent in Control 2 than in the sepsis cohort (Table 1), so that the elevation of renal markers in sepsis cannot be attributed to a higher burden of chronic renal disease. At the same time, creatinine concentrations were markedly elevated in sepsis patients compared to both control cohorts (Figure 5d), demonstrating genuinely impaired renal function at the time of sampling. The observed alterations are instead consistent with sepsis-associated acute kidney injury (SA-AKI), a common manifestation of sepsis [21], in which proteins that accumulate as glomerular filtration declines increase accordingly. Cystatin-C has additionally been reported to be influenced by systemic inflammation independently of glomerular filtration [22], providing a further explanation for its increased abundance. The renal markers within the sepsis core set should therefore not be regarded as artifacts of cohort composition, but as reflecting renal dysfunction as an integral manifestation of the syndrome.

Several proteins related to renal function were also found to be associated with age, consistent with the progressive decline in kidney function over the life course. Beyond these, our analyses identified age-associated proteins involved in extracellular matrix remodeling and the insulin-like growth factor axis. This is in line with previous proteomics studies showing that ageing affects multiple biological pathways and shapes the circulating plasma proteome independently of sepsis [23,24]. Age was, however, not independent of the other covariates, as several age-associated proteins were also linked to renal function (CST3, CFD, COL18A1). This collinearity is biologically expected and illustrates that age-related and renal effects on the plasma proteome cannot be entirely separated.

Another important confounder that we studied was postoperative sterile inflammation. Both the number of differential proteins and the classifier performance were reduced for Control 1 at the postoperative time point, whereas neither was the case for Control 2 (Figure 4a). The clinical routine data indicate that this difference is primarily explained by inflammatory kinetics. The proteome of Control 1 was sampled at the peak of the postoperative acute phase response, while in Control 2 CRP levels were either still rising (day 2) or already falling again (day 5; Figure 5c). The faster CRP increase in Control 1 is consistent with the markedly longer duration of the surgical procedures in this cohort, as the magnitude and kinetics of postoperative inflammation increase with surgical duration and tissue trauma [25–28]. Differences in age and patient numbers between the two control cohorts may have contributed further.

In summary, our study provides a resource of plasma proteome changes associated with sepsis and adds further evidence supporting a set of robustly sepsis-associated proteins. We furthermore characterized the influence of renal conditions, demographic factors and postoperative sterile inflammation on the plasma proteome and provide the corresponding proteins as a reference for future studies (Table 2, Supplementary Tables 13 and 14). The machine learning models complemented the linear modelling, and feature importance analysis identified proteins with discriminative value that were not captured by the linear models, illustrating the added value of combining both approaches.

## Limitations

The encoding of comorbidities as binary categorical variables neglects differences in their severity, thereby representing an approximation of clinical reality. This is particularly relevant for the renal conditions, where the degree of functional impairment rather than the presence of a diagnosis determines the plasma concentrations of the affected proteins. Continuous variables such as creatinine were available only for a subset of the patients and were therefore not included as covariates in the models.

A limitation of the machine learning results is the substantial difference in cohort sizes between the control and sepsis groups. Although Borderline-SMOTE was applied to mitigate class imbalance, synthetic oversampling cannot fully compensate for a limited number of independent observations, potentially limiting model generalizability. Most importantly, no independent validation cohort was available. The reported metrics are cross-validated estimates obtained within a single center and will not directly translate to other patient collectives; the models should therefore be regarded as a proof of concept and as a basis for validation in larger, multicentric cohorts.

The dynamic range of the plasma proteome and the sensitivity of current LC-MS/MS instrumentation limit the detection of low-abundance proteins. In the present study, 1260 proteins were quantified, of which 432 were measured in at least 50 % of the patients. Our observations are therefore restricted to the higher abundant part of the plasma proteome, while low-abundance mediators such as cytokines were not accessible.

## Conclusion

The composition of the plasma proteome is influenced by multiple factors. We demonstrate that renal conditions, demographic factors and sterile inflammation interact with the proteomic alterations induced by sepsis, and that the differences are often quantitative rather than qualitative. These interactions should not be regarded merely as confounding, but as part of the clinical reality of the patients, in which sepsis-induced acute kidney injury, for example, is a common manifestation of the syndrome. Our work shows the importance of considering covariates and sampling time points, and provides a list of proteins that are robustly altered in sepsis. This information supports the future development of novel diagnostic and therapeutic strategies.

## Methods

### Study Population

The study included patients from four prospectively recruited cohorts. Patients with sepsis were derived from the SepsisDataNet.NRW (SDN) study (DRKS00018871) and the KI.SEP study (DRKS00032970), both of which prospectively enrolled adult patients fulfilling the Sepsis-3 criteria and collected biospecimens and clinical data according to predefined study protocols [29]. As SepsisDataNet.NRW recruited at several centers, only patients enrolled at the University Hospital Knappschaft Kliniken Bochum, where KI.SEP and both control cohorts were conducted, were included in the present analysis. Only sepsis patients who survived a minimum of 48 h after enrollment were considered, to exclude the most severe cases, in which extensive organ failure and early mortality might otherwise have biased the comparison with hospital controls.

Two non-septic control cohorts were analyzed to distinguish sepsis-specific alterations from changes associated with surgical stress, hospitalization, and intensive care treatment. Control 1 consisted of hospitalized patients with planned postoperative intensive care or intermediate care monitoring, recruited within the SepsisDataNet.NRW framework; patients fulfilling the Sepsis-3 criteria at study inclusion were excluded. Control 2 consisted of elective surgical patients aged 60 years or older, recruited within the prospective CONFUSED study (DRKS00033854) [30], who underwent surgery under general anesthesia without evidence of sepsis and without a requirement for postoperative intensive or intermediate care monitoring. All participants were at least 18 years of age and provided written informed consent, or consent was obtained from a legal representative where applicable. All studies were approved by the responsible ethics committees (SDN 18-6606-BR and 22-7477, KI.SEP 23-7905, CONFUSED 23-7794 and 2024-082-f-S) and were conducted in accordance with the Declaration of Helsinki.

### Sample Collection and Time Points

Within SepsisDataNet.NRW, blood samples were obtained on the day of study inclusion, within 36 h after sepsis diagnosis. In KI.SEP, samples were collected up to 72 h after sepsis diagnosis and up to 24 h after the start of antibiotic therapy. For both sepsis cohorts, only samples from these baseline time points were used for the present analysis. In both control cohorts, samples were collected immediately before surgery (pre-OP); For Control 1, a second sample was collected on postoperative day 3 (day 3 post-OP), whereas for Control 2, samples were collected on days 2 and 5 post-OP. Patients without an available pre-operative sample were included if at least one postoperative sample was available. In all studies, EDTA blood was used (S-Monovette, Sarstedt, Nümbrecht, Germany), plasma was separated by centrifugation at 1000 × g for 10 min and stored at −80 °C until analysis. C-reactive protein and creatinine concentrations were retrieved from the hospital’s clinical routine laboratory records where available and are presented descriptively.

### Proteomics Sample Preparation

For proteome analysis, plasma samples were processed according to the SP3 protocol with minor modifications as described before [31]. Briefly, 1 µL of each sample was reduced with 20 mM dithiothreitol for 30 min at room temperature and alkylated with 50 mM 2-iodoacetamide for another 30 min in the dark. The reaction was quenched with 50 mM DTT, and a 1:1 mixture of Cytiva Sera-Mag SpeedBeads A and B was added. Acetonitrile was added to a final concentration of 70 %, and the samples were incubated for 20 min before being placed in a magnetic rack. The supernatant was discarded and the beads were washed twice with 70 % ethanol and once with acetonitrile. Proteins were digested overnight at 37 °C with trypsin (1.5 µg in 50 mM ammonium bicarbonate, pH 7.8). Acetonitrile was added to a final concentration of 95 %, the supernatant was discarded, and peptides were eluted with 2 × 25 µL 0.1 % formic acid. Samples were dried and resuspended in 350 µL 0.1 % TFA.

### LC-MS/MS analysis

For LC-MS/MS analysis, 15 µL of the peptide solution was injected onto a DNV PepMap Neo C18 column (15 cm, 75 µm ID, 2 µm particle size) operated at 60 °C in a Vanquish Neo coupled to an Orbitrap Exploris 480 (both Thermo Scientific). Mobile phase A consisted of 0.1 % formic acid; mobile phase B of 80 % acetonitrile and 0.1 % formic acid. Peptides were separated at a flow rate of 350 nL/min using a linear gradient from 4 % to 21 % B over 42 min, followed by an increase to 37 % B over the next 33 min and to 42 % B within a further 4 min. The column was subsequently washed for 15 min from 80 % to 100 % B at a flow rate of 400 nL/min prior to the next injection. For data acquisition, the Exploris 480 was operated in data-independent mode using 38 isolation windows covering 350 to 970 m/z, with one MS1 scan after every 19 MS2 scans. MS1 scans were acquired at a resolution of 120,000 with a maximum injection time of 70 ms, a normalized AGC target of 300 % and an RF lens setting of 55 %. For MS2, a normalized HCD collision energy of 30 %, a resolution of 30,000, a maximum injection time of 80 ms and a normalized AGC target of 3000 % were employed.

### Proteomics Data Analysis

All patient samples were measured in three batches and analyzed separately using DIA-NN (v.2.3.0) searching against the human UniProt/SwissProt database (v.2025_02). The maximum number of missed cleavages was set to two and for all other parameters default settings were used. After raw data processing, a batch normalization was applied as described previously [31]. Quality control was carried out using principal component analysis (PCA) and boxplots (Supplementary Figure 4). Only proteins with at least ten observations throughout the data set were considered for further analyses. Linear models were calculated using the Limma package [32]. Only proteins measured in at least 50 % of the analyzed sub-cohorts were used. False discovery rate (FDR)-adjusted p-values were used throughout the study and for the comparisons of sepsis and control patients, a ratio of means threshold of ≥ 1.5 or ≤ 0.67 was applied. Propensity score matching was carried out using the MatchIt package [33]. Patients with a specific comorbidity were considered as the reference group, and patients without the respective comorbidity were matched in a 1:2 ratio using nearest-neighbor matching on the logit of the propensity score, estimated from a logistic regression model including age, sex, and condition (sepsis or control) as covariates. Age and sex were included as standard demographic covariates and the condition was found to have a major impact on the plasma proteome using linear models (see Figure 2). The resulting matched sub-cohorts were analyzed using t-tests, considering only proteins with at least five observations in each of the compared groups. P-values were corrected according to the Benjamini-Hochberg method. Statistical analyses were carried out using R (v.4.4.3). Proteins found to be differentially abundant were analyzed separately for each of the five comparisons using STRING (v.12.0, string-db.org).

### Machine Learning Models

Four binary classification models were trained, comparing sepsis patients with Control 1 and Control 2 at the pre- and the postoperative time points; no model was trained for Control 2 at day 5 post-OP due to insufficient patient numbers. The minority class, i.e. the respective control cohort, was defined as the positive class. For each classification task, two algorithms were evaluated: logistic regression with Elastic Net regularization (hereafter referred to as Elastic Net) and eXtreme Gradient Boosting (XGBoost) with L1 and L2 regularization. The Elastic Net was selected because it performs well when the number of input features is large relative to the sample size [34], and XGBoost was added as a nonlinear tree-based ensemble method able to capture more complex relationships. Immunoglobulin variable region entries were removed from the feature space, as these correspond to gene segments rather than proteins, making peptide-to-protein assignment ambiguous and complicating biological interpretation. Models were trained by a nested stratified Monte Carlo cross-validation (MCCV) with 100 repetitions. In each repetition, the data set was split into 70 % training and 30 % test data while preserving class ratios, and hyperparameters were tuned within the training set by a randomized grid search using a stratified 3-fold inner cross-validation, optimized for the area under the receiver operating characteristic curve (AUROC) to account for class imbalance (Supplementary Table 2). Within each training fold, missing values (up to 30 % missingness allowed; Supplementary Figure 5) were imputed using scikit-learn’s IterativeImputer implementation of the Multivariate Imputation by Chained Equations (MICE) algorithm (Supplementary Table 3), features were standardized using scikit-learn’s StandardScaler, and the minority class was up-sampled using Borderline-SMOTE. All pre-processing steps were fitted on the training data only and subsequently applied to the test data, which retained the original class distribution, in order to avoid data leakage. Accuracy, sensitivity, specificity, F1-score, Matthew’s correlation coefficient (MCC) and AUROC were determined on the test data of each of the 100 repetitions and averaged for model comparison. To evaluate feature contributions to model decision-making, SHapley Additive exPlanations (SHAP) were computed for each repetition. Features were ranked in descending order of their absolute SHAP value, with rank 0 assigned to the feature with the highest value, and the median rank across the 100 repetitions was used to summarize the contribution of each feature. All libraries and versions are listed in Supplementary Table 4. Model development and evaluation were conducted in accordance with the DOME (Data, Optimization, Model and Evaluation [35] recommendations to promote transparent and reproducible machine learning analyses and were published in the DOME registry with ID zjfm5s62jt.

## Statistics and Reproducibility

The study is an observational analysis of prospectively recruited cohorts. All patients meeting the inclusion criteria with an available plasma sample were analyzed. Each sample represents one biological replicate, no technical replicates were measured. Group assignment followed the clinical condition of the patients. Baseline characteristics were compared using Kruskal-Wallis tests for continuous variables, Fisher exact tests for binary variables and a chi-squared test for infection focus, followed by Dunn’s test or pairwise Fisher exact tests with Bonferroni correction. SOFA score and infection focus were compared between sepsis patients and Control 1 only. The analyses of the protein data are described under Proteomics Data Analysis and Machine Learning Models.

## Supporting information

Supplementary_Table_6_Control_1_pre_OP_multivariate_significant

Supplementary_Table_7_Control_1_day_3_post_OP_multivariate_significant

Supplementary_Table_8_Control_2_pre_OP_multivariate_significant

Supplementary_Table_9_Control_2_day_2_post_OP_multivariate_significant

Supplementary_Table_10_Control_2_day_5_post_OP_multivariate_significant

Supplementary_Table_11_signficant_proteins_propensity_score_matching

Supplementary_Table_12_significant_proteins_sepsis_vs_control

Supplementary_Table_13_significant_proteins_sex

Supplementary_Table_14_significant_proteins_age

Supplementary_Table_16_machine-learning_features_and _meadian_SHAP_ranks

## Acknowledgement

The authors thank the Core Unit Bioinformatics - CUBiMed.RUB at the Faculty of Medicine of the Ruhr-University Bochum for the support with and access to the provided high performance computing facilities. The authors thank Stephanie Adamzik for her excellent support.

## Author Contributions

Conceptualization: MW, AW, TB. Methodology: MW, AW, TB, RG. Investigation: MB, FK, KF, KMW, BW, SB, KR, DZ, BZ. Resources: KMW, BW, SB, KR, DZ, ME. Data curation: KR, DZ, HN, SB, FK. Formal analysis: MW, AW, TB. Writing – original draft: MW, AW, TB. Writing – review and editing: TR, MB, BS, ME. Funding acquisition: AW, FK, BK, ME, MA, HN, TR, BS. Supervision: RG, ME, MA, BS.

## Code availability

The R and Python code used for data processing, statistical analysis, machine learning and figure generation is available from the corresponding author upon reasonable request.

## Data availability

The mass spectrometry proteomics data have been deposited to the ProteomeXchange Consortium via the PRIDE partner repository with the dataset identifier PXD082539.

Reviewer access details:

Log in to the PRIDE website using the following details:

Project accession: PXD082539

Token: cRvSp9HmzBsc

