## Supplementary Material for "Confounder-Adjusted Plasma Proteomics Identify Robust Protein Signatures in Sepsis Across Two Independent Perioperative Control Cohorts"

**Table of Contents**

**Supplementary Tables**

Supplementary Table 1: Distribution of surgical procedures in the two control cohorts.

Supplementary Table 2: Value ranges for the hyperparameters tuned in the inner fold of the Monte Carlo cross-validation.

Supplementary Table 3: Hyperparameters for the MICE imputation of missing values, which was applied for the machine learning.

Supplementary Table 4: Used libraries for pre-processing and training of the machine learning models.

Supplementary Table 5: Completeness of demographic data and comorbidities.

Supplementary Table 6 (Excel file): Control 1, pre-OP – significant proteins from the multivariate model.

Supplementary Table 7 (Excel file): Control 1, day 3 post-OP – significant proteins from the multivariate model.

Supplementary Table 8 (Excel file): Control 2, pre-OP – significant proteins from the multivariate model.

Supplementary Table 9 (Excel file): Control 2, day 2 post-OP – significant proteins from the multivariate model.

Supplementary Table 10 (Excel file): Control 2, day 5 post-OP – significant proteins from the multivariate model.

Supplementary Table 11 (Excel file): Significant proteins identified by propensity score matching.

Supplementary Table 12 (Excel file): Significant proteins, sepsis vs. control.

Supplementary Table 13 (Excel file): Significant proteins associated with sex.

Supplementary Table 14 (Excel file): Significant proteins associated with age.

Supplementary Table 15: Model performance metrics for both timepoints for the Elastic Net model (EN) and XGBoost (XGB).

Supplementary Table 16 (Excel file): Machine learning features and median SHAP ranks.

**Supplementary Figures**

Supplementary Figure 1: Correlation matrix of comorbidities calculated on the sepsis cohort.

Supplementary Figure 2: Sequence coverage of COL18A1.

Supplementary Figure 3: Confusion matrices for the Elastic Net models.

Supplementary Figure 4: Effect of batch normalization on the plasma proteome data set.

Supplementary Figure 4: Binned missingness of feature values.

Supplementary Figure 6: Sensitivity analysis excluding sepsis patients with kidney transplantation or dialysis dependency.

**Supplementary Tables:**

**Supplementary Table 1:** Distribution of surgical procedures in the two control cohorts.

| Type of surgery | Control cohort 1, n/N (%) | Control cohort 2, n/N (%) | p-value |
| --- | --- | --- | --- |
| Joint replacement surgery | 0/71 (0.0%) | 42/73 (57.5%) | <0.001 |
| Surgical fracture fixation | 1/71 (1.4%) | 2/73 (2.7%) | 1.000 |
| Visceral surgery | 23/71 (32.4%) | 5/73 (6.8%) | <0.001 |
| Spinal surgery | 1/71 (1.4%) | 12/73 (16.4%) | 0.002 |
| Vascular surgery | 4/71 (5.6%) | 0/73 (0.0%) | 0.057 |
| Oral and maxillofacial surgery | 12/71 (16.9%) | 0/73 (0.0%) | <0.001 |
| Neck dissection with flap reconstruction | 3/71 (4.2%) | 0/73 (0.0%) | 0.117 |
| Other surgical procedures | 0/71 (0.0%) | 12/73 (16.4%) | <0.001 |
| Cranial neurosurgery* | 27/71 (38.0%) | 0 |  |

Data are presented as n/N (%). *Patients undergoing cranial neurosurgery were not included in control cohort 2 according to the study protocol.

**Supplementary Table 2:** Value ranges for the hyperparameters tuned in the inner fold of the Monte Carlo cross-validation.

| **Algorithm** | **Hyperparameter** | **Value Range** |
| --- | --- | --- |
| xgboost | tree_method | Exact |
|  | gamma | 0.01-0.4 |
|  | n_estimators | 500-1000 |
|  | lambda | 0.1-0.9 |
|  | alpha | 0.1-0.8 |
|  | max_depth | 3-30 |
| Logistic regression | C | 0.001-10 |
|  | solver | Saga |
|  | max_iter | 100-10000 |
|  | L1_ratio | 0.1-0.9 |
| Random Forest | Min_samples_leaf | 3-10 |
|  | Ccp_alpha | 0.001-0.1 |
|  | Oob_score | True |
|  | Class_weight | Balanced |
|  | Max_depth | 6-40 |
|  | Min_samples_split | 6-15 |
|  | Max_features | Log2, sqrt |
|  | N_estimators | 1000 |

**Supplementary Table 3:** Hyperparameters for the MICE imputation of missing values, which was applied for the machine learning.

| **Hyperparameter** | **Value** |
| --- | --- |
| Random_state | None |
| N_nearest_features | 30 |
| Tol | 0.001 |
| Max_iter | 10 |
| Imputation_order | ascending |
| Initial_strategy | mean |

**Supplementary Table 4:** Used libraries for pre-processing and training of the machine learning models.

| Library | Version |
| --- | --- |
| Pandas | 3.0.2 |
| Scikit-learn | 1.8.0 |
| Python | 3.14.4 |
| XGBoost | 3.2.0 |
| Imbalanced-learn (Borderline-SMOTE) | 0.14.1 |
| SHAP | 0.51.0 |

**Supplementary Table 5:** Completeness of demographic data and comorbidities. Values represent numbers of missing values.

|  | **Control 1** | **Control 2** | **Sepsis** |
| --- | --- | --- | --- |
| **Age** | 0 | 0 | 0 |
| **Sex** | 0 | **2** | 0 |
| **BMI** | 0 | **2** | **64** |
| **SOFA Score** | **21** | **75** | 0 |
| **Alcohol** | 0 | 0 | 0 |
| **COPD** | 0 | 0 | 0 |
| **Other lung disease** | 0 | 0 | 0 |
| **Hypertension** | 0 | 0 | 0 |
| **CKD** | 0 | 0 | 0 |
| **Diabetes** | 0 | 0 | 0 |
| **Obesity** | 0 | 0 | **3** |
| **Cardiovasc** | 0 | 0 | **1** |
| **Malignancy** | 0 | 0 | **1** |
| **Nicotine** | 0 | 0 | 0 |
| **Dialysis** | 0 | 0 | **2** |
| **Transplant** | 0 | 0 | 0 |

**Supplementary Table 15:** Model performance metrics for both timepoints for the Elastic Net model (EN) and XGBoost (XGB). MCC stands for Matthew Correlation Coefficient and AUROC for area under the receiver operating curve. Bold marks values with the highest sensitivity for each classifier.

|  | Pre-OP | | | | Post-OP | | | |
| --- | --- | --- | --- | --- | --- | --- | --- | --- |
|  | Control 1 | | Control 2 | | Control 1 | | Control 2 | |
|  | EN | XGB | EN | XGB | EN | XGB | EN | XGB |
| Accuracy | 0.95 | 0.94 | 0.95 | 0.94 | 0.86 | 0.90 | 0.98 | 0.95 |
| Sensitivity | **0.88** | 0.84 | **0.89** | 0.81 | 0.58 | **0.61** | **0.86** | 0.56 |
| Specificity | 0.96 | 0.97 | 0.96 | 0.97 | 0.90 | 0.94 | 0.99 | 0.99 |
| F1-Score | 0.85 | 0.84 | 0.83 | 0.81 | 0.52 | 0.62 | 0.89 | 0.65 |
| MCC | 0.82 | 0.81 | 0.80 | 0.78 | 0.44 | 0.57 | 0.88 | 0.65 |
| AUROC | 0.96 | 0.97 | 0.98 | 0.97 | 0.83 | 0.89 | 0.99 | 0.96 |

**Supplementary Figures**

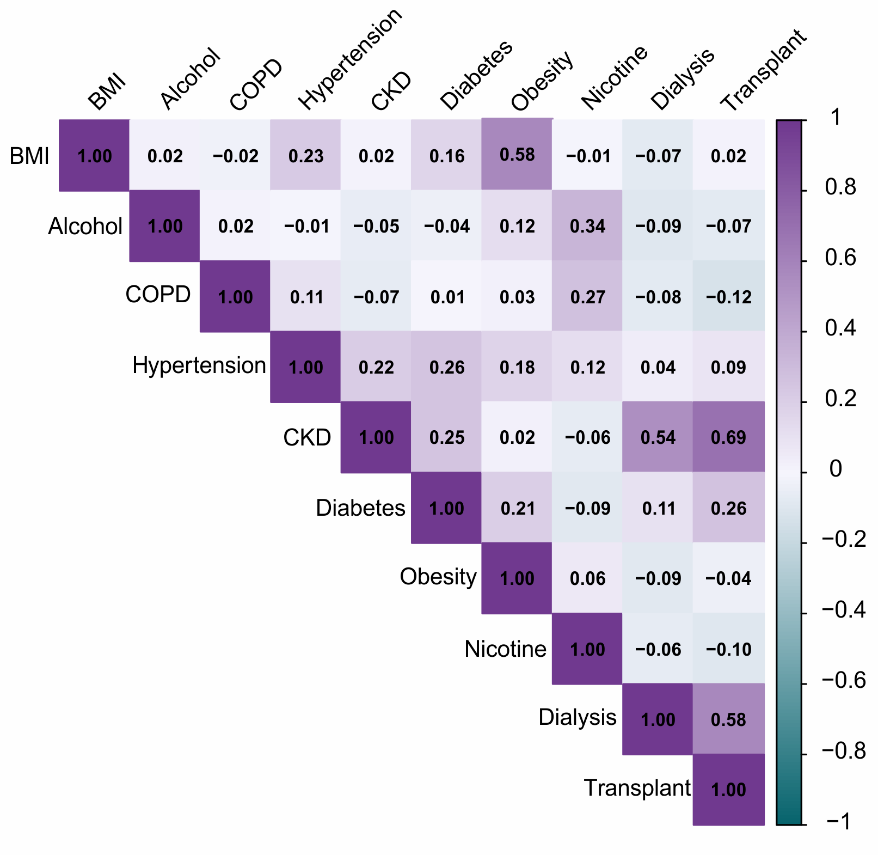

**Supplementary Figure 1: Correlation matrix of comorbidities calculated on the sepsis cohort.**

Pearson correlation matrix for all comorbidities as well as age, sex and BMI (calculated on the sepsis cohort; only terms with at least one correlation > |0.25| displayed).

>sp|P39060|COIA1_HUMAN Collagen alpha-1(XVIII) chain OS=Homo sapiens OX=9606 GN=COL18A1 PE=1 SV=5

MAPYPCGCHILLLLFCCLAAARANLLNLNWLWFNNEDTSHAATTIPEPQGPLPVQPTADT

TTHVTPRNGSTEPATAPGSPEPPSELLEDGQDTPTSAESPDAPEENIAGVGAEILNVAKG

IRSFVQLWNDTVPTESLARAETLVLETPVGPLALAGPSSTPQENGTTLWPSRGIPSSPGA

HTTEAGTLPAPTPSPPSLGRPWAPLTGPSVPPPSSGRASLSSLLGGAPPWGSLQDPDSQG

LSPAAAAPSQQLQRPDVRLRTPLLHPLVMGSLGKHAAPSAFSSGLPGALSQVAVTTLTRD

SGAWVSHVANSVGPGLANNSALLGADPEAPAGRCLPLPPSLPVCGHLGISRFWLPNHLHH

ESGEQVRAGARAWGGLLQTHCHPFLAWFFCLLLVPPCGSVPPPAPPPCCQFCEALQDACW

SRLGGGRLPVACASLPTQEDGYCVLIGPAAERISEEVGLLQLLGDPPPQQVTQTDDPDVG

LAYVFGPDANSGQVARYHFPSLFFRDFSLLFHIRPATEGPGVLFAITDSAQAMVLLGVKL

SGVQDGHQDISLLYTEPGAGQTHTAASFRLPAFVGQWTHLALSVAGGFVALYVDCEEFQR

MPLARSSRGLELEPGAGLFVAQAGGADPDKFQGVIAELKVRRDPQVSPMHCLDEEGDDSD

GASGDSGSGLGDAR**ELLREETGAALKPR**LPAPPPVTTPPLAGGSSTEDSRSEEVEEQTTV

ASLGAQTLPGSDSVSTWDGSVRTPGGRVKEGGLKGQKGEPGVPGPPGRAGPPGSPCLPGP

PGLPCPVSPLGPAGPALQTVPGPQGPPGPPGRDGTPGRDGEPGDPGEDGKPGDTGPQGFP

GTPGDVGPKGDKGDPGVGERGPPGPQGPPGPPGPSFRHDKLTFIDMEGSGFGGDLEALRG

PRGFPGPPGPPGVPGLPGEPGRFGVNSSDVPGPAGLPGVPGREGPPGFPGLPGPPGPPGR

EGPPGRTGQKGSLGEAGAPGHKGSKGAPGPAGARGESGLAGAPGPAGPPGPPGPPGPPGP

GLPAGFDDMEGSGGPFWSTARSADGPQGPPGLPGLKGDPGVPGLPGAKGEVGADGVPGFP

GLPGREGIAGPQGPKGDRGSRGEKGDPGKDGVGQPGLPGPPGPPGPVVYVSEQDGSVLSV

PGPEGRPGFAGFPGPAGPKGNLGSKGERGSPGPKGEKGEPGSIFSPDGGALGPAQKGAKG

EPGFRGPPGPYGRPGYKGEIGFPGRPGRPGMNGLKGEKGEPGDASLGFGMRGMPGPPGPP

GPPGPPGTPVYDSNVFAESSRPGPPGLPGNQGPPGPKGAKGEVGPPGPPGQFPFDFLQLE

AEMKGEKGDRGDAGQKGERGEPGGGGFFGSSLPGPPGPPGPPGPRGYPGIPGPKGESIRG

QPGPPGPQGPPGIGYEGRQGPPGPPGPPGPPSFPGPHRQTISVPGPPGPPGPPGPPGTMG

ASSGVRLWATRQAMLGQVHEVPEGWLIFVAEQEELYVRVQNGFRKVQLEARTPLPRGTDN

EVAALQPPVVQLHDSNPYPRREHPHPTARPWR**ADDILASPPR**LPEPQPYPGAPHHSSYVH

LRPARPTSPPAHSHR**DFQPVLHLVALNSPLSGGMR**GIRGADFQCFQQAR**AVGLAGTFR**AF

LSSR**LQDLYSIVR**RADRAAVPIVNLKDELLFPSWEALFSGSEGPLKPGAR**IFSFDGKDVL**

**RHPTWPQKSVWHGSDPNGRR**LTESYCETWR**TEAPSATGQASSLLGGR**LLGQSAASCHHAY

IVLCIENSFMTASK

**Supplementary Figure 2: Sequence coverage of COL18A1.**

Primary sequence of human Collagen alpha-1 (XVIII) chain. Yellow highlights mark the sequence of Endostatin. Sequences representing measured peptides are underlined and bold.

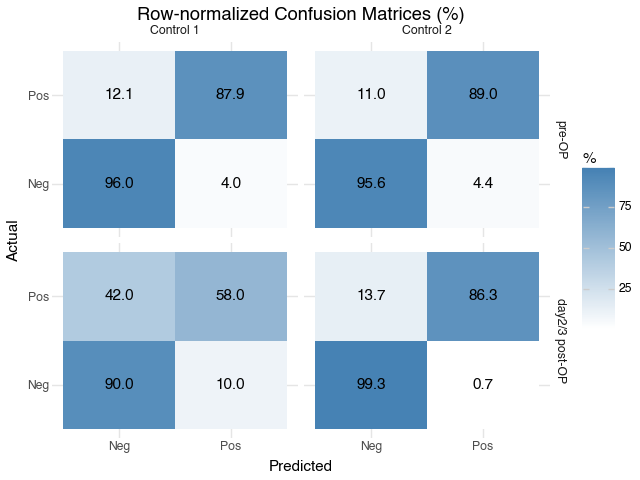

**Supplementary Figure 3:** Confusion matrices for the Elastic Net models. The numbers of true positives, true negatives, false positives and false negatives were also calculated and averaged to create a row-normalized confusion matrix Percentages sum up to 100% row-wise.

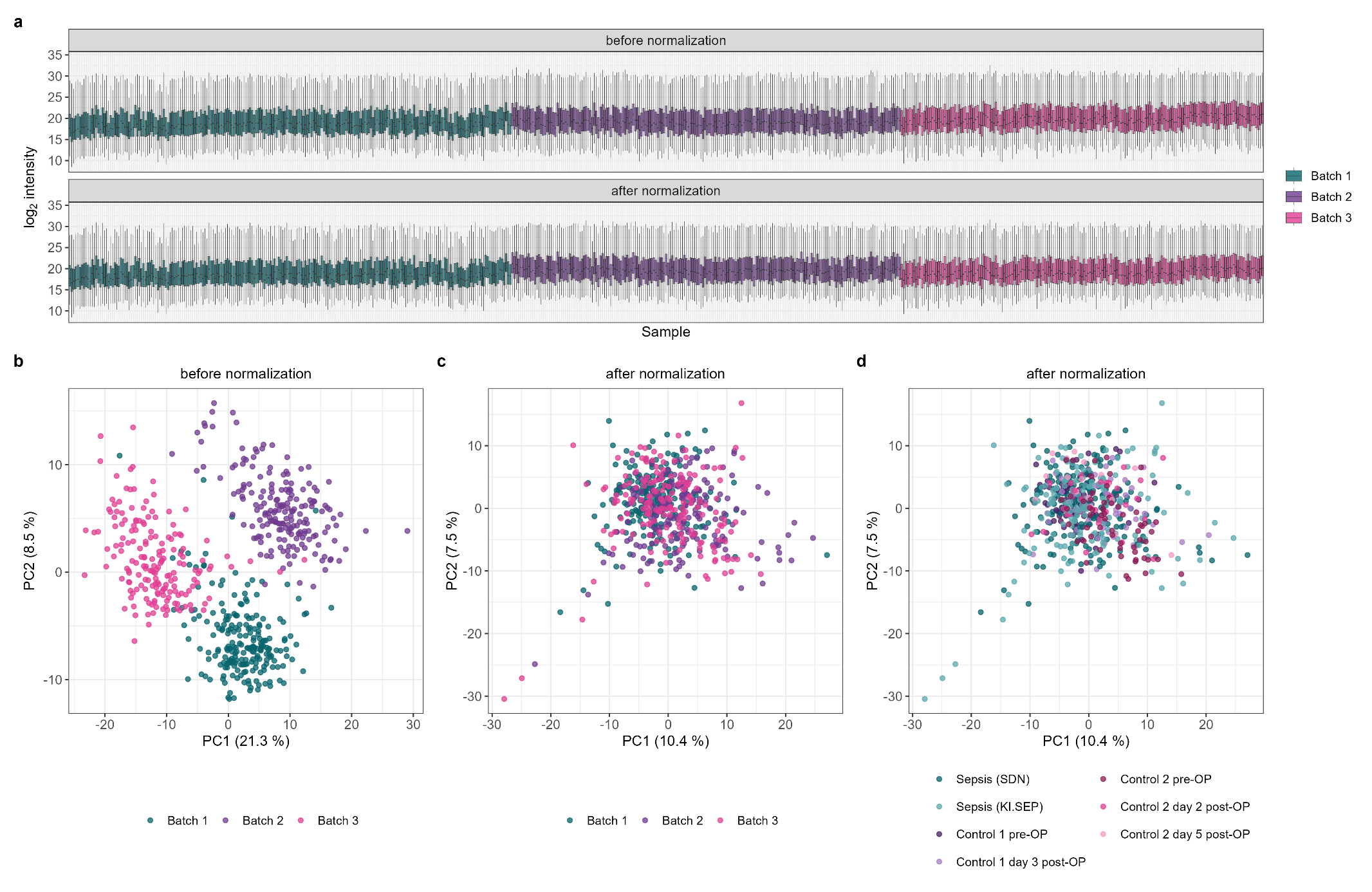

**Supplementary Figure 4: Effect of batch normalization on the plasma proteome data set.**

All samples were measured in three batches on the same instrument and were processed separately in DIA-NN before batch normalization was applied. **a)** Sample-wise distributions of log2 protein intensities before and after normalization. Each box represents one sample, samples are ordered by batch and outliers are not displayed. **b, c)** Principal component analysis of all proteins quantified in at least 50 % of the samples, with remaining missing values replaced by the respective protein median, before **(b)** and after **(c)** normalization, colored by batch. Before normalization, the three batches separated along the first principal component, which accounted for 21.3 % of the total variance. After normalization the batches were no longer separated and the variance explained by the first principal component decreased to 10.4 %. **d)** The same analysis as in **(c)**, colored by study group.

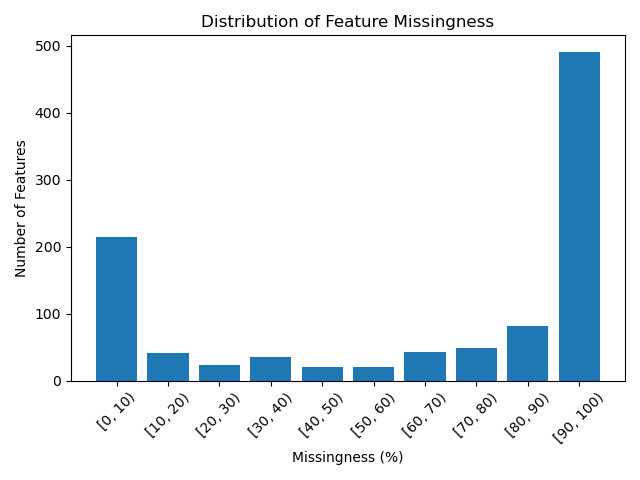

**Supplementary Figure 5: Binned missingness of feature values.**

A cutoff of 30% missing features was chosen for the machine learning models to retain as many features as possible while limiting the number of imputed features.

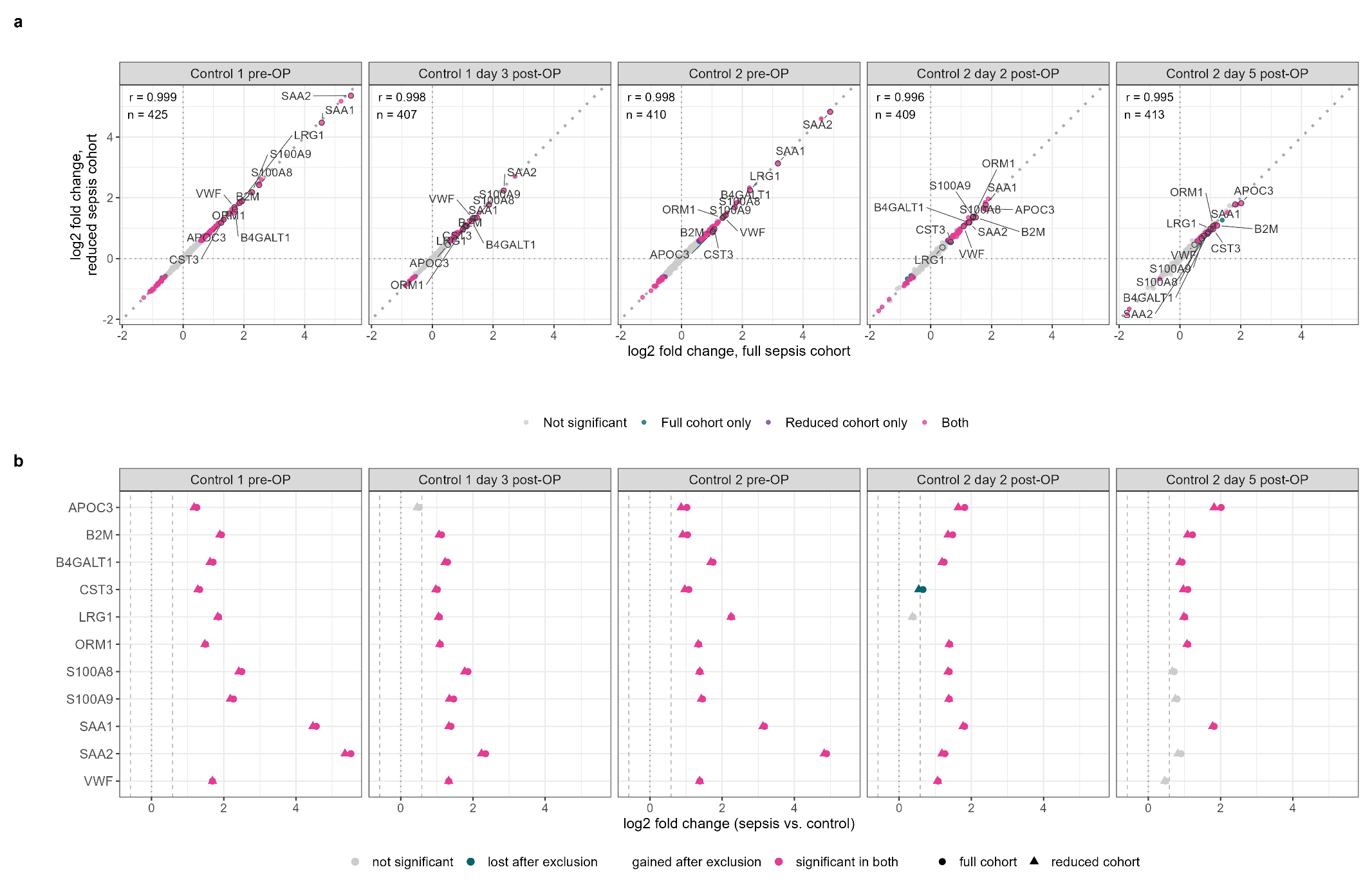

**Supplementary Figure 6: Sensitivity analysis excluding sepsis patients with kidney transplantation or dialysis dependency.** The five multivariate comparisons were repeated after removal of all sepsis patients with a history of kidney transplantation or dialysis dependency (n = 58 of 343), while the control cohorts remained unchanged. **a)** Effect sizes of the condition coefficient (sepsis vs. control) in the full and in the reduced sepsis cohort. Each point represents one protein; the dotted diagonal indicates identical effect sizes. Colours indicate whether a protein met the significance criteria of the main analysis (adjusted p-value < 0.05 and ratio of means ≥ 1.5 or ≤ 0.67) in the full cohort only, in the reduced cohort only, in both, or in neither. Pearson correlation coefficients and the number of compared proteins are given per comparison. The eleven proteins robustly associated with sepsis (Table 3) are circled and labelled with gene names. **b)** Effect sizes of these eleven proteins in both analyses; dashed vertical lines mark the ratio of means thresholds.
